# Elderly acceptance of telemedicine use in Hong Kong during and after the COVID-19 pandemic: a cross-sectional cohort survey

**DOI:** 10.1101/2021.07.15.21260346

**Authors:** Maxwell Chun Yin Choi, Shing Him Chu, Lok Lam Siu, Anakin Gajy Tse, Justin Che Yuen Wu, Hong Fung, Billy Chi Fai Chiu, Vincent Chung Tong Mok

## Abstract

**Background:** Telemedicine services worldwide have experienced an unprecedented boom since the beginning of the COVID-19 pandemic. Multiple studies have noted telemedicine as an effective alternative to traditional face-to-face management of patients. This study provides insight into public perception and impression of telemedicine in Hong Kong, specifically among the elderly who are the most vulnerable to COVID-19.

**Methods:** Face-to-face surveys were conducted on elderly relatives of current medical students at the Chinese University of Hong Kong who were aged ≥ 60 years. The survey included socio-demographic details; past medical history; and concerns towards telemedicine use. Univariate and multivariable regression analyses were conducted to examine statistically significant associations. The primary outcomes are consideration of telemedicine use during: (1) a severe outbreak; and (2) after the COVID-19 pandemic.

**Results:** 109 surveys were conducted. Multivariable regression analyses revealed that expectation of government subsidies for telemedicine services was the strongest common driver, and also the only positive independent predictors of telemedicine use for both during a severe outbreak, as well as after the COVID-19 pandemic. No negative independent predictors of telemedicine use during severe outbreak were found. Negative independent predictors of telemedicine use after the COVID-19 pandemic included old age, and living in the New Territories.

**Conclusions:** Government support such as telemedicine-specific subsidies will be crucial in promoting telemedicine use in Hong Kong both during a severe outbreak and after the current COVID-19 pandemic. Robust dissemination of information regarding the pros and cons of telemedicine towards the public, especially towards the elderly population, is warranted.

## Introduction

The COVID-19 pandemic has led many medical services around the world to experience dwindling numbers, as patients are cancelling clinic appointments or hospital visits out of fear of infection.[1] Telemedicine [2] is defined as: “… the practice of medicine over a distance, in which interventions, diagnoses, therapeutic decisions, and subsequent treatment recommendations are based on patient data, documents and other information transmitted through telecommunication systems.” The ongoing COVID-19 pandemic has subsequently served as a catalyst for the expansion and rising interest in telemedicine services as a potential solution to bridge the gaps in healthcare delivery and minimize the risk of COVID-19 transmission.[3]

Despite the availability of numerous virtual technological solutions, Hong Kong has yet to see significant progress towards widespread implementation and promotion of telemedicine.[4] This raises concerns regarding the ability of hospitals and clinics to provide continued quality care to the masses amid a potential total lock-down situation if COVID-19 cases surge to new heights. Elucidating the reasoning behind telemedicine’s utilization in Hong Kong from the patients’ perspective will not only help identify current deficiencies of the healthcare system, but also facilitate a smoother implementation of telemedicine in the future.

The main objective of the study is to examine the main concerns of elderly towards using telemedicine and subsequently evaluate telemedicine use under 2 different hypothetical scenarios ---(1) during a severe outbreak under a lockdown scenario; (2) after the COVID-19 pandemic. In this study, “severe outbreak” refers to a sudden increase in disease frequency within a limited geographical area which necessitates public health interventions (e.g. a government-imposed lockdown involving temporary restrictions on travel and social interactions, along with quarantine measures),[5] whereas “after COVID-19 pandemic” refers to the likely new norm (i.e. COVID-19 becoming an endemic with regular vaccines required, and societies adapting to the seasonal deaths and illnesses they bring without requiring lockdowns, masks or social distancing).[6]

Our study focused primarily on the elderly population due to age being the greatest risk for severe COVID-19 infections,[7,8] hence telemedicine will likely have the greatest benefit among the elderly population.

## Methods

### Study design and participants

The study consisted of an online survey done by an elderly cohort in Hong Kong SAR, China. The survey was conducted from 8 October to 15 November 2020, between the 3rd and 4th wave of local COVID-19 outbreaks.[9,10] Medical student helpers from the Chinese University of Hong Kong were recruited to assist in data collection from elderly population among their families. Under this consideration, we deemed random visits of elderly face-to-face to have a high risk of infection.[11] Therefore, our study opted to interview the close relatives of our medical students who live within the same household as a safer and more effective way for medical students and participants to reduce the risk of infection.[11]

A total of 59 medical student helpers (Supplementary Data 2) were recruited in September 2020. To ensure the survey protocol (Supplementary Data 1) would be standardized, a mandatory virtual training course was held on 28 September 2020 via Zoom, including a detailed written interview guide (Supplementary Data 3) to supplement the medical student helpers in delivering the survey.

### Procedures

The survey is targeted at the elderly population in Hong Kong, defined by aging 60 or above.[12] For the elderly who were unable to express themselves due to health reasons, their closest caretakers were substituted to represent the elderly for survey completion,[13] as they accompanied the elderly to clinical appointments and they both shared highly similar visiting experiences. The surveys were to be completed and submitted online; consent was obtained from the participants before the start of each survey, which would be administered by a trained medical student helper.

The survey consisted of multiple-choice questions aiming to address five major aspects that could influence elderly perception of telemedicine: (1) socio-demographic characteristics, including age, gender, education level, number of cohabitants in the same household, employment status and residential area; (2) past medical history, including suffering from the types of chronic illness, frequency and difficulty of visiting regular doctors in public and private sectors respectively, the number and types of prescribed medication, as well as enrollment status in private health insurance plans; (3) domestic support of telemedicine usage, including the availability of digital devices and internet access; (4) consideration of telemedicine use in 2 different scenarios (i.e. during severe outbreak, after pandemic); and (5) telemedicine-associated values, concerns and expectations (e.g. concerns about reduced effectiveness and lower satisfaction).

The survey can be found in Supplementary Data 1.

### Statistical analysis

Data analysis was carried out using IBM Statistical Package for Social Sciences (SPSS) Version 26. Our cohort survey responses were first thematically grouped into four main categories: (1) demographics; (2) home characteristics; (3) medical-related features; and (4) telemedicine-related features. The 2 main primary outcome variables in our study were dichotomous variables on consideration of telemedicine use: (1) during severe outbreak and (2) after the COVID-19 pandemic.

Multivariable regression analysis was conducted by selecting only statistically significant (i.e. *P*<.05) variables according to results from univariate regression analysis. In summary, we performed four independent binary logistic regression analyses for the aforementioned 2 main primary outcome variables using both univariate and multivariable analyses.

## Results

### Cohort characteristics

Among the 109 respondents interviewed by 59 medical student helpers, 93.6% (N=102/109) consisted of elderly aged ≥ 60 years, while 6.4% (N=7/109) were caretakers who acted as surrogate interviewees on behalf of their elderly.

The detailed characteristics of our cohort is shown in Table 1. The average age of our respondents was 73±10 years, with most of them being female (57.8%, n=63/109) and having a secondary level of education or above (68.8%, n=75/109). In terms of the respondent’s household characteristics, 44% (n=48/109) of them lived in New Territories, with each household’s average size being 2.9±1.5 members. Although most had both access to the Internet (93.6%, n=102/109) and digital devices (91.7%, n=100/109), none had used telemedicine before, so none were able to comment on what type of telemedicine they would prefer.

**Table 1:** Characterization of elderly cohort (N=109)

|  |  |
| --- | --- |
|  | **Total (n=109)** |
| **Demographics** |  |
| Elderly respondent | 102 (93.6%) |
| Age, years | 72.7 (10) |
| Gender (Female) | 63 (57.8%) |
| Education |  |
| *Primary or below* | 34 (31.2%) |
| *Secondary* | 48 (44%) |
| *Tertiary* | 27 (24.8%) |
| **Home characteristics** |  |
| District |  |
| *New Territories* | 48 (44%) |
| *Kowloon* | 38 (34.9%) |
| *Hong Kong Island* | 23 (21.1%) |
| Number of Household members | 2.9 (1.5) |
| Internet access | 102 (93.6%) |
| Digital devices | 100 (91.7%) |
| No previous use of telemedicine | 109 (100%) |
| **Medical-related features (i.e. past medical history)** |  |
| Types of diseases |  |
| *Cardiovascular* | 57 (52.3%) |
| *Metabolic/endocrine* | 32 (29.4%) |
| *Musculoskeletal* | 22 (20.2%) |
| Number of different types of drugs taken regularly | 2 (2) |
| Number of doctors/specialties consulted regularly |  |
| *At Public sector* |  |
| *0 doctor* | 44 (40.4%) |
| *1-3 doctors* | 59 (54.1%) |
| *4-6 doctors* | 6 (5.5%) |
| *At Private sector* |  |
| *0 doctor* | 45 (41.3%) |
| *1-3 doctors* | 63 (57.8%) |
| *4-6 doctors* | 1 (0.9%) |
| Having a private medical insurance coverage | 45 (41.3%) |
| **Telemedicine-related features** |  |
| Values avoiding hospital/clinic environment due to fear of infection | 73 (67%) |
| Expects that government subsidies will increase likelihood of telemedicine use | 70 (64.2%) |
| Concerned about reduced effectiveness and lower satisfaction | 50 (45.9%) |
| Values shorter waiting time | 33 (30.3%) |
| Values maintaining doctor-patient relationship | 19 (17.4%) |
| Values are presented as mean (standard deviation) / count (percentage) as appropriate. |  |

The survey also collected the detailed past medical and drug history in our cohort. In terms of disease epidemiology, the main chronic disease was cardiovascular disease (52.3%, n=57/109), followed by metabolic/endocrine (29.4%, n=32/109) and then musculoskeletal disease (20.2%, n=22/109), whereas the mean number of drugs taken was 2±2. Most of our respondents regularly consulted 1-3 doctors in both the public (54.1%, n=59/109) and private sector (57.8%, n=63/109), yet less than half have private medical insurance coverage (41.3%, n=45/109). As for telemedicine-related responses, the majority of our cohort values avoiding hospital or clinic environments due to the fear of infection (67%, n=73/109), and also expects government subsidies [14] would increase their likelihood of utilizing telemedicine (64.2%, n=70/109).

On the other hand, almost half of the respondents are concerned about the reduced effectiveness and lower satisfaction levels gained from telemedicine use (45.9%, n=50/109); however, less than half valued the maintenance of a doctor-patient relationship (17.4%, n=19/109) or shorter waiting time (30.3%, n=33/109) with telemedicine use.

### Factors affecting telemedicine use during severe COVID-19 outbreak

Multivariable regression analysis (Table 2) showed that expectation of government subsidies to increase telemedicine services was the only positive independent predictors for telemedicine usage during severe outbreak (aOR= 5.043, 95% CI 1.353-18.795; *P*=.016). No negative independent predictors for considering telemedicine use during severe outbreak were found.

**Table 2:** Factors affecting telemedicine use during a severe outbreak

|  | CONSIDER telemedicine during severe outbreak |  |  |  |
| --- | --- | --- | --- | --- |
|  | Univariate |  | Multivariable |  |
|  | OR (95% CI) | *P value* | aOR (95% CI) | *P value* |
| **Demographics** |  |  |  |  |
| Age, years | 0.943 (0.897 - 0.992) | **.023** | 0.985 (0.92-1.056) | .673 |
| Gender (Female) | 0.895 (0.333-2.404) | .825 |  |  |
| Education |  | .471 |  |  |
| *Primary or below* | Reference |  |  |  |
| *Secondary* | 1.333 (0.456-3.902) | 0.6 |  |  |
| *Tertiary* | 2.462 (0.584-10.371) | .22 |  |  |
| **Home characteristics** |  |  |  |  |
| District |  | .268 |  |  |
| *New Territories* | 1.474 (0.372-5.836) | .581 |  |  |
| *Kowloon* | 0.589 (1.161-2.518) | .425 |  |  |
| *Hong Kong Island* | Reference |  |  |  |
| Number of Household members | 1.419 (0.989-2.035) | .058 |  |  |
| Internet access | 1.867 (0.335-10.393) | .476 |  |  |
| Digital devices | 1.302 (0.249-6.792) | .755 |  |  |
| **Medical related features** |  |  |  |  |
| Types of diseases |  |  |  |  |
| *Cardiovascular* | 0.682 (0.254-1.828) | .447 |  |  |
| *Metabolic/endocrine* | 1.306 (0.431-3.958) | .636 |  |  |
| *Musculoskeletal* | 0.511 (0.17-1.535) | .232 |  |  |
| Number of different types of drugs taken regularly | 0.79 (0.626-0.997) | **.047** | 0.958 (0.668-1.374) | .814 |
| Number of doctors/specialties consulted regularly |  |  |  |  |
| At Public sector |  | **.006** |  | .475 |
| *0 doctor* | Reference |  | Reference |  |
| *1-3 doctors* | 0.436<br>(0.129-1.476) | .182 | 1.303<br>(0.263-6.463) | .746 |
| *4-6 doctors* | 0.02<br>(0.002-0.216) | **.001** | 0.252<br>(0.011-5.912) | .392 |
| At Private sector |  | **.044** |  | .132 |
| *0 doctor* | Reference |  | Reference |  |
| *1-3 doctors* | 3.859<br>(1.337-11.137) | **.013** | 4.257<br>(1.039-17.441) | .044 |
| *4-6 doctors* | / | / | / | 1 |
| Having a private medical insurance coverage | 3.417<br>(1.058-11.033) | **.04** | 1.637<br>(0.338-7.925) | .54 |
| **Telemedicine-related features** |  |  |  |  |
| Concerned about reduced effectiveness and lower satisfaction | 0.291<br>(0.102-0.828) | **.021** | 0.424<br>(0.106-1.691) | .224 |
| Values shorter waiting time | 2.881<br>(0.782-10.612) | .112 |  |  |
| Values maintaining doctor-patient relationship | 0.155<br>(0.052-0.465) | **.001** | 0.225<br>(0.046-1.113) | .067 |
| Values avoiding hospital/clinic environment due to fear of infection | 4.062<br>(1.480-11.150) | **.007** |  |  |
| Expects that government subsidies will increase likelihood of telemedicine use | 8.125<br>(2.664-24.781) | **<.001** | 5.043<br>(1.353-18.795) | **.016** |

### Factors affecting telemedicine use after the COVID-19 pandemic

Multivariable regression analysis (Table 3) showed that expectation of government subsidies for telemedicine services was also the strongest common driver and the only positive independent predictor for telemedicine use after the pandemic (aOR=5.498, 95% CI 1.470-20.572; *P*=.011). However, we discovered 2 negative independent predictors for telemedicine use after the pandemic: old age (aOR=0.913; 95% CI 0.846-0.985; *P*=.019); and living in New Territories (aOR=0.102; 95% CI 0.024-0.433; *P*=.002).

**Table 3:** Factors affecting telemedicine use after the COVID-19 pandemic

|  | CONSIDER telemedicine after pandemic |  |  |  |
| --- | --- | --- | --- | --- |
|  | Univariate |  | Multivariable |  |
|  | OR (95% CI) | P value | aOR (95% CI) | P value |
| **Demographics** |  |  |  |  |
| Age, years | 0.907 (0.864-0.951) | **<0.001** | 0.913<br>(0.846-0.985) | **.019** |
| Gender (Female) | 0.396 (0.18- 0.872) | **.021** | 0.674<br>(0.231-1.966) | .47 |
| Education |  | **.019** |  | .36 |
| *Primary or below* | Reference |  | Reference |  |
| *Secondary* | 1.2 (0.464--3.106) | .707 | 0.609<br>(0.165-2.253) | .458 |
| *Tertiary* | 4.08 (1.393-11.947) | **.010** | 1.641<br>(0.315-8.539) | .556 |
| Home characteristics |  |  |  |  |
| District |  | **.027** |  | **.005** |
| *New Territories* | 0.239 (0.083-0.684) | **.008** | 0.102<br>(0.024-0.433) | **.002** |
| *Kowloon* | 0.468 (0.163-1.345) | .158 | 0.526<br>(0.131-2.114) | .366 |
| *Hong Kong Island* | Reference |  | Reference |  |
| Number of Household members | 1.146 (0.891- 1.475) | .289 |  |  |
| Internet access | 4.2(0.488-36.180) | .191 |  |  |
| Digital devices | 1.333 (0.315-5.642) | .696 |  |  |
| **Medical related features** |  |  |  |  |
| Types of diseases |  |  |  |  |
| *Cardiovascular* | 0.361 (0.163-0.799) | **.012** | 0.968<br>(0.321-2.920) | .954 |
| *Metabolic/endocrine* | 1.544 (0.670-3.560) | .308 |  |  |
| *Musculoskeletal* | 0.849 (0.322-2.236) | .74 |  |  |
| Number of different types of drugs | 1.239 (0.992 – 1.549) | .059 |  |  |
| Number of doctors/specialties consulted regularly |  |  |  |  |
| *At Public sector* |  | .082 |  |  |
| *0 doctor* | Reference |  |  |  |
| *1-3 doctors* | 0.401 (0.178-0.902) | **.027** |  |  |
| *4-6 doctors* | 0.457 (0.076-2.755) | .393 |  |  |
| *At Private sector* |  | .151 |  |  |
| *0 doctor* | Reference |  |  |  |
| *1-3 doctors* | 2.238 (0.993-5.042) | .052 |  |  |
| *4-6 doctors* | / | / |  |  |
| Having a private medical insurance coverage | 4.5 (1.978-10.239) | **<0.001** | 1.772<br>(0.533-5.886) | .35 |
| **Telemedicine related features** |  |  |  |  |
| Concerned about reduced effectiveness and lower satisfaction | 0.402 (0.181-0.896) | **.026** | 0.495<br>(0.17-1.446) | .199 |
| Values shorter waiting time | 1.429 (0.624-3.272) | .399 |  |  |
| Values maintaining doctor-patient relationship | 0.234 (0.064-0.861) | **.029** | 0.367<br>(0.073-1.842) | .223 |
| Values avoiding hospital/clinic environment due to fear of infection | 2.145 (0.905-5.083) | .083 |  |  |
| Expects that government subsidies will increase likelihood of telemedicine use | 3.875 (1.564-9.603) | **.003** | 5.498<br>(1.470-20.572) | **.011** |

## Discussion

### Interpretation of Results

Multivariable regression analysis revealed no negative independent predictors for discouraging telemedicine usage during severe outbreaks. This may be explained by the fear of COVID-19 infection within the Hong Kong population, prompting citizens to avoid public transport and practice social distancing.[15] Having previously experienced the SARS epidemic of March 2003, many Hong Kong citizens are still fearful of unknown infectious diseases.[16] Given that telemedicine carries no risk of infection compared to traditional face-to-face consultations,[17] it certainly has value under an epidemic or pandemic scenario; however, it has been shown in literature that telemedicine would underperform in hands-on procedures, such as physical examination or postoperative care.[18] Nonetheless, with rapid technological advancements, these limitations may soon be overcome. Hence, it is therefore reasonable to infer that the nature and benefits of telemedicine potentially outweigh its limitations under a severe outbreak scenario, including an epidemic or even pandemic.

For both “severe outbreak” and “after pandemic” scenarios, expectation of government subsidies for telemedicine services was the strongest common driver of telemedicine use. It was also the only statistically significant positive independent predictors of considering telemedicine use for after the COVID-19 pandemic ends. For example, the Elderly Health Care Voucher Scheme was launched in Hong Kong in 2009, aiming to provide financial incentives to elderly when seeking medical services in the private sector so as to alleviate stress placed on the public healthcare system. So far, this scheme has contributed to positive encouragement of telemedicine usage in the private sector.[14] Therefore, we propose that telemedicine-specific subsidies should be made available to the elderly population in the pandemic to encourage use of telemedicine services.

Furthermore, the role of governmental support in promoting telemedicine use should be emphasized and expanded. For instance, subsidies can be provided to Hong Kong’s elderly population to purchase essential digital devices for telemedicine consultations, such as webcams and remote monitoring devices. Indeed, taking Australia as an example, government support in healthcare by relaxing insurance reimbursement restraints has been fundamental towards the growth of telemedicine use in Australia.[19] Moreover, public education on telemedicine and digital health in general should be executed to clarify patient misconceptions and expectations towards telemedicine. For example, it is indispensable to emphasize that telemedicine use does not mean that patients should default on follow-up. We strongly recommend further education on the format (i.e. video call, using digital health applications etc.), effectiveness (i.e. limited physical examination) and other expectations of telemedicine as these were the most important concerns in our elderly cohort.

There were 2 statistically significant negative independent predictors for considering telemedicine use after the COVID-19 pandemic: old age and living in New Territories. For older generations, weak technological competency is undoubtedly a challenge to overcome when adapting to an entirely new consultation practice. Elderly often struggle with unfamiliar technology which may ultimately deter many from utilizing telemedicine. To help elderly patients adopt new technologies, telemedicine systems should be designed to be as accessible as possible.[20] For example, easy to navigate interfaces, simple instructions with larger display fonts, may help increase the elderly population’s willingness to use telemedicine for follow-up of chronic illness after the pandemic.

When considering elderly who live in the New Territories, a relatively more rural part of Hong Kong, digital infrastructure necessary to carry out telemedicine services may not be as adequate when compared to Hong Kong Island and Kowloon. New Territories also houses the greatest number of poverty hotspots in Hong Kong, which may correlate low socio-economic status and education with lower healthcare utilization.[21] The same study also correlates poverty rate with access to hospitals, with New Territories generally having the lowest level of access to hospitals among all districts; yet telemedicine may in fact be the most suitable for their residents as it provides healthcare services considering the distance to hospitals and clinics.[21] In short, telemedicine accessibility in Hong Kong remains a major problem warranting further study.

### Strengths

This is the first study in Hong Kong which has holistically examined the concerns towards telemedicine implementation from the perspective of a cohort of elderly citizens, both during and after the pandemic. Utilizing telemedicine as a novel approach to managing patients may serve as a crucial component of an effective healthcare system during a pandemic, and could even be implemented as a formidable adjunct to in person consultations in post-pandemic clinical practice.[22]

Additionally, the mandatory training course for students and detailed breakdown of each question ensured adequate quality control and a full understanding of telemedicine prior to carrying out the survey (Supplementary Data 1). The training course also endeavored to homogenize the delivery of surveys, thereby minimizing potential confirmation or observer bias resulting from unstandardized survey delivery styles among different helpers. The interview guide (Supplementary Data 3) was explicitly introduced in the course, which accounted for the detailed rationale of our study, as well as key points to take note of for each survey question. Further information regarding current background and applications of telemedicine, expectations of our medical student helpers, as well as proper communication and history-taking skills were presented; the training course was recorded for absentees who were unable to attend the live investigator meeting.

### Limitations

There is a possibility of selection bias and inter-observer bias, since the survey only includes the responses of family members of medical students. However, this is compensated by our training course for standardization. Retrospectively, the study results have demonstrated no definite tendency for our respondents to favor telemedicine, suggesting that any potential selection bias is essentially negligible.

Additionally, this survey involved a relatively small sample size due to pandemic-associated social distancing restrictions; For instance, our cohort did not involve residents living in outlying islands in Hong Kong. Located remotely and having low access to hospitals,[21] these areas may showcase a greater need for telemedicine. Therefore, caution should be taken before generalization to other populations outside of Asia. Furthermore, this study was performed before the formal introduction of COVID-19 vaccination programs, which has been shown to influence the general public’s attitude towards health-seeking behavior.[23] Therefore, this study may not be fully representative of the most updated pandemic situation in Hong Kong.

Moreover, in our survey, we directly correlated and likened the scenario of a severe outbreak in Hong Kong to that of the grave lockdown situation in Wuhan due to having no prior lockdown experience in Hong Kong at the time of survey completion. As such, the gravity of what constitutes a ‘severe’ outbreak may vary depending on the individual’s subjective interpretation. Notably, as none of our survey participants have actually utilized telemedicine in the past due to its relative novelty in Hong Kong, the intention and willingness to utilise telemedicine can only approximate the actual use of telemedicine. In fact, we predict that the future implementation and functionality of telemedicine in Hong Kong will ultimately depend on alterations in health-seeking behavior of the general public both during and after the COVID-19 pandemic.

### Future studies

This study was primarily targeted at the elderly. Future studies should be done to investigate the responses of adults, adolescents and children to telemedicine as well. The perspectives of caretakers and elderly themselves could also be compared in future studies on a larger scale to account for possibly contrasting concerns among different stakeholders.

## Conclusion

In summary, this study has examined the main concerns of elderly towards using telemedicine both during a severe outbreak under a lockdown scenario, and after the COVID-19 pandemic. Our results have highlighted government support as a crucial driver of telemedicine usage in Hong Kong under both scenarios. To overcome the hesitance towards telemedicine use stemming from technological inconveniences due to old age and geographical location, telemedicine-specific subsidies and public education will be essential to spearhead telemedicine use after the pandemic.

Government support, in the form of telemedicine-specific subsidies, will be a crucial driver of telemedicine use in Hong Kong for both during severe outbreak and arguably even more so after the COVID-19 pandemic. In order to sustain telemedicine practice after the pandemic, design of telemedicine systems should be as elderly-friendly as possible. However, telemedicine should be used together with traditional face-to-face consultation and should not be considered as a total replacement.

**What is already known**

Telemedicine has been largely spearheaded during the ongoing COVID-19 pandemic in many countries to minimise the risk of infection. Elderly will benefit greatly from utilizing telemedicine. However, Hong Kong has seen minimal progress towards implementation of telemedicine. This study aims to uncover the concerns of elderly in Hong Kong towards telemedicine to better safeguard their health in both the current and future pandemics.

**What this study adds**

No factors discouraged telemedicine use during a severe pandemic, while old age and living in New Territories discouraged telemedicine use after the pandemic. Government subsidies encouraged telemedicine use in both scenarios. Therefore, government support such as telemedicine-specific subsidies and public education will be crucial towards encouraging telemedicine use in Hong Kong.

## Data Availability

After publication, access to supplementary data files will be made available upon reasonable request to the corresponding author. De-identified participant data will be provided after approval by the authors.

## Acknowledgements

We expressed our gratitude to the Division of Neurology, Department of Medicine and Therapeutics, The Chinese University of Hong Kong, and CUHK Medical Centre for supporting our research, Brian Yiu for his assistance in biostatistics and to all medical student helpers for helping to conduct the face-to-face surveys.

## Conflicts of interest

### Author contributions

All authors contributed equally.

## Ethics Statements

Ethics approval for the study was obtained from The Joint Chinese University of Hong Kong – New Territories East Cluster Clinical Research Ethics Committee (reference number: 2020.536). The study conforms to the Declaration of Helsinki.

